# S4D-ECG: A shallow state-of-the-art model for cardiac abnormality classification

**DOI:** 10.1101/2023.06.30.23292069

**Authors:** Zhaojing Huang, Luis Fernando Herbozo Contrera, Leping Yu, Nhan Duy Truong, Armin Nikpour, Omid Kavehei

## Abstract

An algorithm for processing raw 12-lead ECG data has been developed and validated in this study that is based on the S4D model. Among the notable features of this algorithm is its strong resilience to noise, enabling the algorithm to achieve an average F1-score of 81.2% and an AUROC of 95.5%. It is characterized by the elimination of pre-processing features as well as the availability of a low-complexity architecture that makes it suitable for implementation on numerous computing devices because it is easily implementable. Consequently, this algorithm exhibits considerable potential for practical applications in analyzing real-world ECG data.

## 1 Introduction

The Electrocardiogram (ECG) serves as a non-invasive diagnostic modality for monitoring the electrical activity of the human heart. This diagnostic technique entails strategically placing electrodes on the patient’s body to capture the ECG signal. The widely recognized and standardized method for electrode placement in ECG monitoring is the 12-lead ECG, which incorporates six chest and six limb leads. These leads enable the measurement of electrical heart activities along horizontal and vertical planes [15]. Various AI models have emerged as potential tools for diagnosing abnormalities in the current landscape. Research has introduced a deep neural network specifically designed to analyse electrocardiogram (ECG) data, achieving remarkable performance with F1-scores exceeding 80% and specificity exceeding 99% [16]. The proposed model utilized a residual network architecture to process ECG signals effectively [16]. Petmezas et al. [15] developed a CNN-LSTM model with a sensitivity of 97.87% and specificity of 99.29%. In another available work, a deep neural network integrated with a genetic algorithm and machine learning techniques is devised, which results in enhanced performance for abnormality diagnosis [8]. This approach achieved an average accuracy of 94% and an impressive F1-score of 95.3% [8]. Furthermore, Alfaras et al. [2] proposed an Echo State Networks model based on single-lead ECG signals, which achieved a sensitivity of 92.7% and a positive predictive value of 75.1%. Yildirim et al. [20] contributed to the field by developing a 1D convolutional neural network specifically designed for processing 10-second-long ECG data[20]. The model achieved an impressive accuracy of 91.33%.

### 1.1 Background

Before this work, several other research papers have utilized Structured State Space Sequence (S4) Models for ECG analysis. Notably, Mehari and Strodthoff [12] developed an S4 model that achieved a supervised performance of 0.9417 ± 0.0016 when tested with the PTB-XL dataset. This significant achievement demonstrates the effectiveness and potential of S4 models in accurately analyzing and classifying ECG signals. In addition, Alcaraz and Strodthoff [1] developed an S4 model as a diffusion-based approach for the generation of ECG data. Their model conditioned a diverse set of 71 ECG statements in a multilabel setting, representing an unprecedented degree of complexity [1]. This work showcases the versatility of S4 models and their ability to handle complex ECG data generation tasks.

This study utilises the Diagonal State Space Sequence (S4D) algorithm to demonstrate its out-of-sample generalization, robustness against noise, and its efficacy and significance in processing hospital-grade 12-lead electrocardiogram (ECG) data without preprocessing. We found that although a higher number of layers can help improve the model’s performance, the actual increase is not significant. Therefore, a four-layer model is chosen.

The current study primarily focuses on 12-lead ECG data, which is commonly used in the intensive care unit and clinical settings for comprehensive cardiac monitoring. However, considering the need for device applications and daily monitoring scenarios, the development of algorithms for 1-lead ECG will be necessary in the future.

Figure 1 proposes a visionary device for early detection and timely intervention of abnormalities. Healthcare professionals can benefit from this technology by focusing on specific points where the abnormality is detected, reducing their workload. Integrating AI in portable devices also enables remote monitoring and telemedicine, which is particularly beneficial for individuals in remote areas.

**Figure 1:**
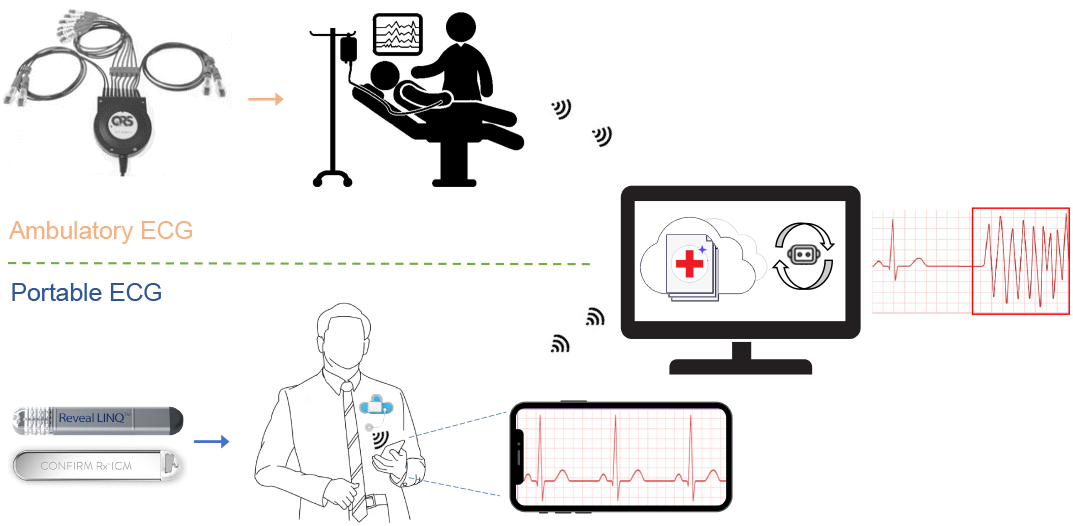
The proposed AI-based abnormality detection model can be implemented in hospital settings and portable devices for assistance with the detection of abnormal heart rhythms. For example, Universal ECG™ by VectraCor (top) and Medtronic’s LINQ or Abbott’s Confirm Rx™ Insertable Cardiac Monitor (ICM) devices (bottom).

## 2 Prerequisite

### 2.1 Structured State Space Sequence (S4) Model

The Structured State Space sequence model (S4) is a deep learning model specifically designed to process long data sequences. Recognizing long-range dependencies is critical when dealing with complex data sets exhibiting intricate patterns and relationships [6]. It has been demonstrated to offer a compelling alternative to more traditional sequence models like RNNs, CNNs, or Transformers.

The continuous-time state-space model converts a one-dimensional input signal *u*(*t*) into a latent state *x*(*t*) of higher dimensions before projecting it onto a one-dimensional output signal *y*(*t*).

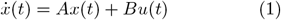

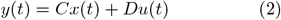

Given matrices *A, B, C*, and *D* of appropriate sizes, the state-space model (SSM) can be discretized using various techniques to model sequences with a fixed step size Δ. One commonly used technique is the zero-order hold method. By employing this method, we can derive a simple linear recurrence relationship as follows:

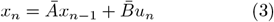

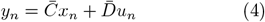

Here, 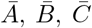, and 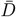 can be calculated as functions of *A, B, C, D*, and Δ.

### 2.2 Diagonal State Space Sequence (S4D) Model

The S4 model is designed to capture the structure and dependencies in time series data using a structured state space model formulation. It incorporates a parameterization technique known as the diagonal plus low-rank (DPLR) matrix to represent the state matrix, allowing it to capture hierarchical dependencies efficiently.

On the other hand, the S4D model simplifies the parameterization by using a diagonal state space model formulation [7]. This means that the state matrix is represented as a diagonal matrix without the low-rank term. This simplification reduces the computational complexity and makes the model easier to implement and understand. By removing the low-rank term from the parameterization, the S4D model achieves a simpler mathematical representation compared to the S4 model.

## 3 Datasets

The proposed model in this study was evaluated using two different datasets: the 12-lead ECG dataset and the Telehealth Network Minas Gerais (TNMG) dataset [16]. The 12-lead ECG dataset, also known as the CPSC dataset, was created as part of The China Physiological Signal Challenge 2018 [11]. This dataset is specifically designed for the automatic identification of rhythm and morphology abnormalities in 12-lead ECGs. It serves as the primary dataset for training the model in this study.

To assess the generalization performance of the model, the TNMG dataset was used as a separate test dataset. The TNMG dataset is a collection of ECG data from the Telehealth Network Minas Gerais project, which focuses on remote healthcare delivery in the Minas Gerais region of Brazil. This dataset provides an opportunity to evaluate how well the model performs on real-world data outside of the CPSC dataset.

By training the model on the CPSC dataset and testing it on the TNMG dataset, the researchers aimed to assess the model’s ability to generalize and perform well on unseen data. This evaluation helps determine the model’s effectiveness and reliability in real-world scenarios.

### 3.1 CPSC

The CPSC dataset has been compiled to encompass 12-lead electrocardiograms (ECGs) that have been recorded at a sampling rate of 500 Hz. The dataset is notable for its collection of ECGs from 6,877 patients exhibiting an array of cardiovascular conditions and common rhythms, all of which have been expertly labeled [11]. To effectively assess the model’s performance, we trained it using 8 distinct abnormalities chosen from the dataset. The table below provides further elaboration on the 8 selected abnormalities.

In the selection process, any data with empty readings were also removed from the dataset. Consequently, the dataset contains 6,877 unique tracings. The data were reformatted into 4,096 readings, and any additional readings were excluded from the dataset. The specifics of the refined dataset are outlined in Figure 2a).

**Figure 2:**
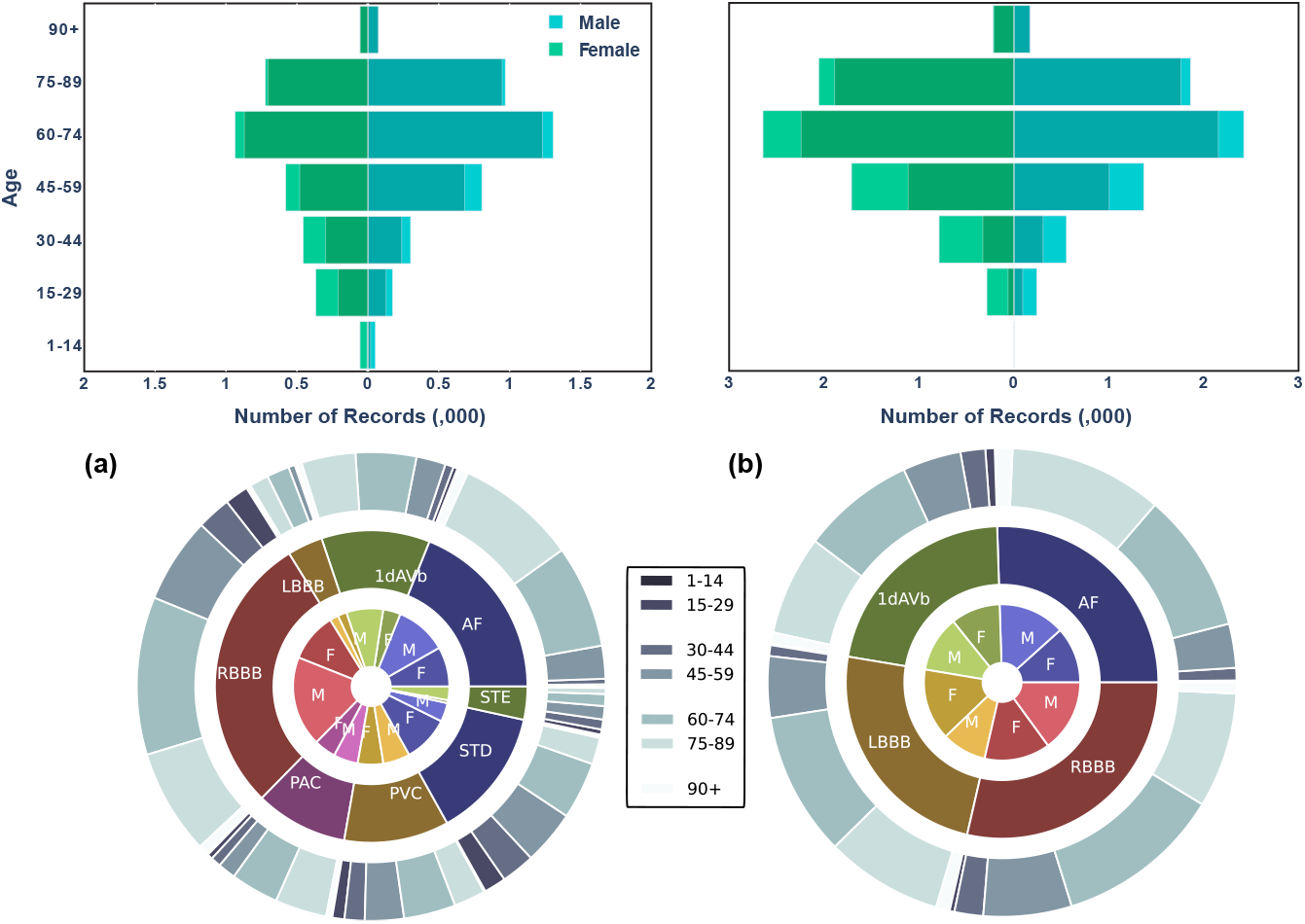
For a), the top left chart of the CPSC dataset displays age and gender categorization, where a darker bar represents the data related to abnormality. In the bottom left chart, the central doughnut graph presents the prevalence of various types of abnormality. The inner layer of the doughnut illustrates the distribution by gender, while the outer layer represents the distribution by age group. For b), the top right chart of the TNMG subset displays age and gender categorization, where a darker bar represents the data related to abnormality. In the bottom right chart, the central doughnut graph presents the prevalence of various types of abnormality. The inner layer of the doughnut illustrates the distribution by gender, while the outer layer represents the distribution by age group.

The figure reveals more male than female patients, indicating a gender imbalance in the dataset. Additionally, most individuals in the dataset are patients rather than non-patients. The age distribution of the patients follows a similar trend to that of the general population, with a larger proportion of individuals in the older age groups. However, there is a slight imbalance when examining the distribution of abnormality. Particularly, the instances of LBBB and STE are relatively fewer than the other types of abnormalities in the dataset.

### 3.2 TNMG

The TNMG dataset comprises 2,322,513 labelled 12-lead electrocardiogram (ECG) data featuring six distinct types of abnormalities [16], sampled at a rate of 400 Hz. To ensure conformity with the CPSC dataset, the data is resampled to 500 Hz. Of the six abnormalities present, four types overlap with those in the CPSC dataset, namely Atrial Fibrillation (AF), First Degree Atrioventricular Block (1dAVb), Left Bundle Branch Block (LBBB), and Right Bundle Branch Block (RBBB). The remaining two abnormalities, Sinus Bradycardia (SB) and Sinus Tachycardia (ST), do not overlap with the CPSC dataset. To evaluate the generalization performance of the trained model in a balanced dataset, 3000 samples were randomly selected from each of the four overlapping abnormalities, with an additional 3000 samples featuring no abnormality [10]. Consequently, the expected size of the sampled dataset is 15,000. However, given that some of the samples exhibit 2 or more abnormality, the total size of the dataset is 14,332. Fig. 2b) below illustrates the dataset’s characteristics.

The dataset readings are standardized to a fixed length of 4096, with any additional readings beyond this length being removed from the data. This standardization process ensures consistency in the data’s structure and facilitates its analysis and modelling. The resampled dataset exhibits a balanced gender distribution, with a roughly equal proportion of males and females. The age distribution in the resampled dataset also follows the general trend of the population, with a higher proportion of individuals in the older age range. Additionally, the sampling process employed in creating the resampled dataset has resulted in a balanced distribution of the different abnormalities.

## 4 Method

We aim to develop a simple model that efficiently handles raw data and employs a shallow architecture suitable for implementation on hardware devices. Even though our tests have been conducted on 12-lead ECG, the low complexity architecture and lack of need for preprocessing features mean that the solution can enable simpler front-end electronics (e.g. reduced need for filtering) and highly accurate and portable heart monitoring system relying on a smaller number of leads in future. This includes hardware devices with limited processing capabilities, such as embedded systems, IoT devices, or edge computing platforms. It ensures optimal performance while utilizing the available hardware resources effectively. The model will prioritize simplicity and computational efficiency, enabling the direct processing of raw data without extensive filtering or feature engineering. This approach reduces latency and facilitates faster analysis of real-time data.

### 4.1 Diagonal State Space Model

The model integrates Diagonal State Space Models, a widely recognized framework [7], renowned for its effectiveness in capturing and understanding the dynamics and interactions within time series data. By incorporating the principles of Diagonal State Space Models into its design, the model demonstrates a dedicated focus on processing time sequence data.

Diagonal State Space Models provide a robust framework for modelling and analyzing time series data. They allow for the representation of both observed and latent variables, which are unobserved variables that influence the behaviour of the observed variables. Using convolutional operations of S4D on signals instead of traditional convolutions on images empowers the model to effectively capture the underlying dynamics, dependencies, and interactions inherent within the data. The proposed model architecture, as shown in Figure 3, was evaluated through experiments to determine the optimal number of S4D layers for achieving the best results. In contrast, a simple dense layer was employed as the model’s decoder to streamline the architecture.

**Figure 3:**
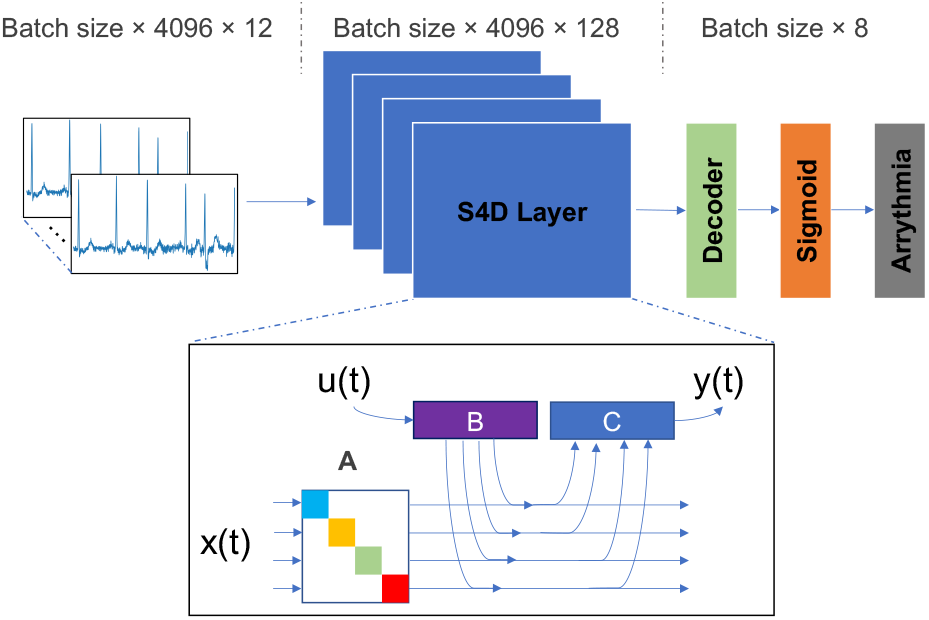
We propose a simple S4D model with raw input data, stacked S4D layers, and a decoder at the end, utilizing the sigmoid function to convert the output into abnormality types. In the S4D layers, the input signal *u*(*t*) is mapped to the output *y*(*t*) through a latent state *x*(*t*) using matrices *A, B*, and *C*.

### 4.2 Evaluation Metrics

The performance metrics used in this paper are: Accuracy 5: The proportion of correct predictions out of the total number of predictions [3].

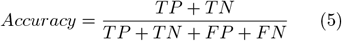

F1-score 8: The harmonic mean of precision 7 and recall 6, which balances the trade-off between precision and recall.

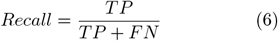

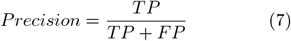

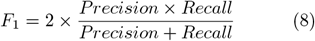

The area under the Receiver Operating Characteristic Curve (AUROC): A measure of the model’s ability to distinguish between positive and negative cases at different threshold values.

The Area Under the Precision-Recall Curve (AUPRC) is a performance metric for binary classification models. It quantifies the overall quality of the model by measuring the area under the precision-recall curve. A higher AUPRC value indicates better model performance, with 1.0 representing a perfect classifier.

These metrics are commonly used in evaluating the performance of machine learning models for binary classification tasks such as abnormality detection.

## 5 Experiment

### 5.1 Training

To determine the optimal number of layers for S4D, we conducted experiments with different layer configurations. We tested S4D models with 2, 4, 6, 8, and 10 layers while keeping other parameters constant. The key parameters used in these experiments included a learning rate of 0.001, 200 epochs, a batch size of 32, a dimension of 128 for the S4D layer output, and a binary cross-entropy loss function.

The CPSC dataset was employed to train the models, and the objective was to observe the variations in behaviour resulting from different architectural designs based on the number of layers used in S4D.

### 5.2 In-sample Test

A subset of 500 data points was chosen from the CPSC dataset specifically for testing the model’s performance. These data points were not utilized during the model’s training phase but were exclusively reserved for evaluating the model’s performance. Using this independent test dataset, we can assess how well the model generalizes to new, unseen data and obtain a more accurate understanding of its overall performance.

Based on the performance results obtained from the testing phase, the best-performing model is selected. The selection process involves comparing the performance metrics of different models and choosing the one that demonstrates the highest level of performance.

### 5.3 Out-of-sample Test

The selected model will be rigorously tested on the TMNG dataset, consisting of 14,332 ECG data points. This dataset is specifically chosen to evaluate the model’s performance in real-world scenarios. The model’s ability to generalize and accurately predict unseen data will be assessed through an out-of-sample test. The predictions will be compared to known values to measure performance metrics. The results will be recorded and analyzed to understand the model’s strengths and limitations. This evaluation will inform decisions regarding the model’s effectiveness and potential refinements for improved ECG analysis.

### 5.4 Robustness Test

During the robustness testing phase, the model will undergo evaluation by randomly emptying leads from the 12-lead ECG data. The aim is to assess the model’s ability to maintain performance even when certain leads are missing. The test will be conducted by progressively emptying different numbers of channels, specifically 2, 4, 6, 8, and 10 leads.

Performance metrics will be used to evaluate the model’s robustness in handling missing leads. By comparing the model’s performance across the different scenarios, we can gain insights into its ability to maintain accurate predictions and interpret ECG data effectively, even when specific leads are absent.

The results of the robustness test will be carefully evaluated to determine how well the model handles the missing lead scenarios. This assessment will help identify potential limitations or areas for improvement in the model’s robustness. By understanding the model’s performance under these conditions, we can make informed decisions about its suitability for practical applications where missing leads may occur in real-world ECG data.

## 6 Result

The results of the experiment are delineated in the ensuing two subsections. The initial subsection showcases the discoveries of the proposed model, whereas the subsequent subsection scrutinizes the proficiency of the proposed model in the context of generalization.

### 6.1 Pre-processing of data

The results obtained from both denoised and non-denoised data, as demonstrated in Table 2, indicate that the proposed model can handle noise effectively for abnormality identification. The denoising techniques employed in the study included a bandpass filter with a low-cut frequency of 0.5 and a high-cut frequency of 40, with an order of 4. Additionally, wavelet denoising was utilized with the ‘db4’ wavelet, a decomposition level of 8, a cutoff frequency of 0.1 Hz, and a filter order of 6. These findings suggest that the step of data preprocessing may not be necessary, as the model performs well even without extensive pre-processing techniques.

**Table 1:** Classifications of abnormalities in CPSC dataset.

| Type | Acronyms | Description |
| --- | --- | --- |
| First Degree Atrioventricular Block | 1dAVb | The presence of a PR interval exceeding 0.2 seconds serves as an indication [13]. |
| Atrial Fibrillation | AF | The most frequently observed form of abnormality is identified by a fast and irregular heartbeat [17]. |
| Left Bundle Branch Block | LBBB | The sequence of activation in the right ventricle preceding the left ventricle leads to alterations in the workload, mechanics, and perfusion of the left ventricle [14]. |
| Right Bundle Branch Block | RBBB | When there is a disruption in signal transmission within the His-Purkinje system, it can result in delayed depolarization in the right ventricle. This condition can then affect the electrical activity of both ventricles, leading to a heart condition [19]. |
| Premature Atrial Contraction | PAC | Abnormal heart rhythm characterized by early depolarizations originating in the atria, the upper chambers of the heart [9]. |
| Premature Ventricular Contraction | PVC | Abnormal heart rhythms that occur when there is an early electrical impulse originating from the ventricles, the lower chambers of the heart [4]. |
| ST-segment Depression | STD | Specific abnormality observed on an electrocardiogram (ECG) where the ST segment, which is a portion of the ECG waveform, is lower in amplitude or depressed below the baseline [18]. |
| ST-segment Elevated | STE | Abnormality observed on an electrocardiogram (ECG) where the ST segment, a portion of the ECG waveform, is elevated above the baseline [5]. |

**Table 2:** Model performance on denoised & raw data.

| Data | F1 | AUROC | AUPRC |
| --- | --- | --- | --- |
| Pre-processed | 84% | 98% | 91% |
| Raw | 85% | 97% | 91% |

### 6.2 S4D Model

The present study involved training the proposed model on the previously introduced CPSC dataset, with a total of 200 epochs. Throughout the training process, both the training and validation loss and accuracy were recorded per epoch. The ensuing figures visually represent the recorded loss and accuracy metrics for both the training and validation sets during the entire training procedure. Furthermore, the S4D model underwent various modifications with different layers, and the obtained results are concisely summarized.

As depicted in Figure 4a), the training loss exhibits a steady decline throughout the training process while the training accuracy increases and stabilizes. While higher layer configurations in the S4D model generally yield improved performance during training, the corresponding increase in accuracy is not significant beyond four layers.

**Figure 4:**
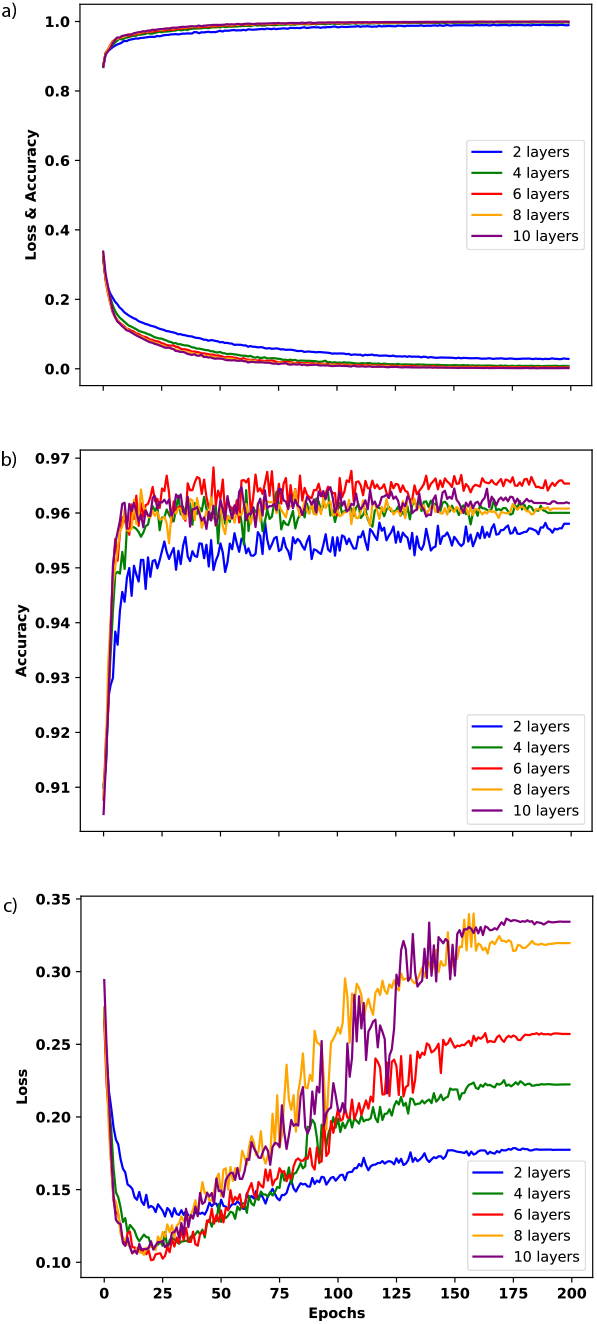
a) illustrates the loss and accuracy metrics performance during the training process of the S4D model with varying layers. b) illustrates the accuracy metric performance during the validation process of the S4D model with varying layers. c) illustrates the loss metric performance during the validation process of the S4D model with varying layers.

Throughout the training process, the validation accuracy generally displays an increasing trend. However, notable fluctuations can also be observed until approximately 175 epochs, after which the accuracy stabilizes. Additionally, Figure 4b) reveals that, apart from the simplest 2-layer model, the number of layers in the S4D model does not significantly affect the validation accuracy.

Figure 4c) depicts the validation loss metrics during the training process. It can be observed that the validation loss exhibits a general trend of initially decreasing and then increasing, with notable fluctuations until approximately 175 epochs, after which it stabilizes. Moreover, it is evident that, for the S4D model, the number of layers directly influences the final validation loss, with higher layer configurations resulting in higher stabilised losses.

After analyzing the behaviour of various models during the training process, we have chosen a four-layer model as the foundation for further analysis. This particular model has exhibited desirable characteristics, including high accuracy and stability in both training and validation performance. Furthermore, the four-layer model offers a simpler architecture compared to other alternatives, as it has a shallow structure. This decision was made based on the model’s performance and the desire for a more straightforward and interpretable model design.

According to Table 3, the 4-layer model performs effectively in detecting abnormality, with a weighted average F1-score of 0.844 and an AUROC of 0.976. Notably, the model demonstrates strong performance in abnormality identifications.

**Table 3:** Model performance on CPSC test set.

| Class | F1 | AUROC | AUPRC | Accuracy |
| --- | --- | --- | --- | --- |
| 1dAVb | 85.2% | 96.2% | 92.9% | 96.8% |
| AF | 92.0% | 98.2% | 95.8% | 97.2% |
| LBBB | 96.8% | 100.0% | 100.0% | 99.8% |
| RBBB | 91.0% | 98.8% | 96.9% | 95.0% |
| PAC | 57.5% | 92.0% | 61.1% | 93.8% |
| PVC | 74.4% | 92.6% | 76.3% | 95.6% |
| STD | 83.8% | 97.7% | 91.3% | 95.2% |
| STE | 66.7% | 97.8% | 65.4% | 98.4% |
| Weighted Average | 84.4% | 97.6% | 91.1% | 96.5% |

**Table 4:** Model generalisation performance on TNMG subset.

| Class | F1 | AUROC | AUPRC | Accuracy |
| --- | --- | --- | --- | --- |
| 1dAVb | 74.8% | 93.0% | 83.4% | 90.9% |
| AF | 71.1% | 94.4% | 87.6% | 88.9% |
| LBBB | 90.6% | 97.9% | 95.8% | 95.9% |
| RBBB | 87.2% | 96.4% | 92.3% | 92.9% |
| Weighted Average | 81.2% | 95.5% | 89.6% | 92.2% |

### 6.3 Model Generalisation

In order to evaluate the generalization performance of the selected model, we utilized the models trained on CPSC to make predictions on the TNMG subset data, which was previously introduced in the data section. Although the original TNMG dataset consists of six distinct types of abnormalities, only four of them overlap with the CPSC dataset. To ensure a balanced dataset, the TNMG subset was specifically created for this purpose. By testing the chosen model on this subset, we can gain insights into its ability to generalize beyond the training data.

Tables 6.2 and 6.3 demonstrate that the model generalizes well, achieving an average F1-score of 0.812 and AUROC of 0.955. This finding illustrates that with a balanced dataset, the model’s generalization capability can achieve high levels of accuracy.

### 6.4 Model Robustness

Based on the promising generalization results achieved with the model, we further evaluated the model on the TNMG dataset under conditions where some channels of the 12-lead ECG data were randomly emptied.

As shown in Figure 5, the model’s performance decreases as the number of emptied leads increases. However, even with half of the 12-lead emptied, the model remains highly robust, with an F1-score above 0.7. These results suggest that the model is capable of handling missing channel data and maintaining its accuracy, indicating its strong robustness.

**Figure 5:**
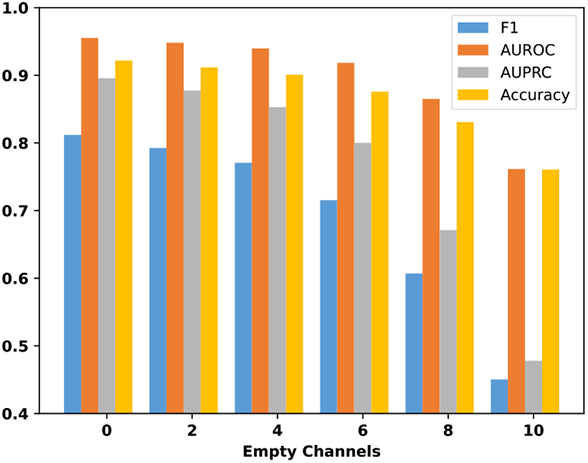
Model performance with a varied number of empty leads of the 12-lead ECG data.

## 7 Discussion

The experiment results indicate that the proposed model for abnormality identification is effective and robust in handling noise. The model demonstrated good performance on both denoised and raw data. The lack of need for a preprocessing or feature extraction step means the system is simpler, architecturally less complex and capable of being deployed in resource-constrained applications such as for portable daily use.

The four-layer model effectively detected abnormalities on the CPSC test set, achieving a weighted average F1-score of 0.844 and an AUROC of 0.976. These results indicate that the model can accurately identify various types of abnormality. Notably, the model achieved particularly strong performance in detecting LBBB, with outstanding F1-score, AUROC, and AUPRC. These highlight the model’s ability to classify specific abnormality classes accurately.

To assess the generalization capability of the selected model, it was tested (inference only) on an out-of-distribution independent dataset, the TNMG dataset from Brazil. The model exhibited good generalization performance with an average F1-score of 0.812 and an AUROC of 0.955. These findings indicate that the model can effectively generalize beyond the training data and accurately classify abnormalities in previously unseen datasets, which is not a common practice observed in many of the publications in this domain and can reliably constitute a pseudo-perspective method of testing and predicting algorithms’ clinical performance.

Furthermore, the model’s robustness was verified by randomly emptying channels of the 12-lead ECG data in the TNMG dataset. The results from this test showed that even with half of the leads emptied, the model remained highly robust and kept an F1 score above 0.7. This demonstrates the model’s ability to handle missing channels, which is a real-world scenario where a complete 12-lead ECG may experience a sudden or persistent lack of data from one or more leads.

Overall, the proposed model exhibits strong performance in abnormality identification, with good generalization capability and robustness to missing channel data. The model’s simplicity and interpretability, combined with its high accuracy, make it a promising tool for abnormality detection in clinical settings. Future research is under development, focusing on evaluating the model’s performance on more extensive and diverse datasets relative to cardiologist performance to be used as an AI-assistant tool in annotating or interpreting ECG data. We hope that the model can be part of a more significant deployment to portable devices and reduces the resources required (memory and energy needs) for near-sensor data analysis.

## 8 Conclusion

This research article introduces a demonstrated algorithm that utilizes the S4 model to process raw and long sequences of ECG data. The algorithm exhibits robustness against noise and demonstrates high efficacy. It is trained on the CPSC dataset (from China) and generalized (inference-only) on the TNMG dataset (from Brazil) subset to test its generalization capabilities. The results showcase an impressive average F1-score of 0.812, providing substantial evidence of the algorithm’s robustness in classification. Furthermore, the AUROC metric achieves a score of 0.955, indicating the model’s notable discriminative ability.

One noteworthy advantage of this algorithm is its ability to handle noise without needing a preprocessing or feature extraction step. We have demonstrated that the accuracy of the results remains unchanged, with or without these steps, affirming the algorithm’s capability to handle noise effectively. As a result, the algorithm offers a low-complexity architecture, enabling deployment on various devices such as embedded systems, medical IoT devices, or near-sensor computing platforms for hospitals and home use. This versatility allows the algorithm to perform even in resource-constrained environments (i.e. hardware), making it a valuable solution for a wide range of real-world applications.

## Data Availability

All data produced in the present study are available upon reasonable request to the authors

## 9 Acknowledgement

Zhaojing Huang would like to acknowledge the support of the Research Training Program (RTP) provided by the Australian Government.

Luis Fernando Herbozo Contreras would like to acknowledge the partial support of the Faculty of Engineering Research Scholarship provided by the University of Sydney.

## 10 Code Availability

To obtain the code used in this paper, simply request it from the corresponding author, but be aware of any conditions or restrictions on its use.

## 11 Data Availability

This paper employs a publicly accessible CPSC dataset, in addition to the TNMG dataset, which can be obtained by requesting access from the data owner. The dataset is not publicly available, but access can be granted upon request with approval from the data owner.

## 12 Declarations

The authors have no conflicts of interest, whether financial or non-financial, to disclose.

